# Causal links between mitophagy and Alzheimer’s disease: a Mendelian randomization study

**DOI:** 10.1101/2024.10.01.24314751

**Authors:** Weijian Zhang, Wenjia Chen, Zhendong Guo, Hui Li

## Abstract

**Background:** Emerging evidence highlights the dysregulation of mitophagy, the process of clearing damaged mitochondria, as a potential contributor to Alzheimer’s disease (AD) pathology. However, the precise mechanisms linking mitophagy to AD remain poorly understood. This study utilized summary-data-based Mendelian Randomization (SMR) combined with multi-omics data to explore the causal relationships between them, and to uncover potential epigenetic mechanisms of gene regulation.

**Methods:** Mitophagy-related genes were identified through the integration of three databases, and transcriptomic data of AD patients were obtained from the Gene Expression Omnibus (GEO) database. A meta-analysis was conducted to recognize differentially expressed genes (DEGs) associated with mitophagy in AD. Through SMR tools, genome-wide association study (GWAS) summary data of AD from the GWAS Catalog (n=487,511) were separately integrated with expression quantitative trait loci (eQTLs) and DNA methylation quantitative trait loci (mQTLs) from blood and brain tissues to identify potentially causal genes and methylation sites. The findings from primary analysis were validated with data from the UK Biobank (n=301,478).

**Results:** In total, 111 mitophagy-related genes were found to be differentially expressed in AD. The three-step SMR analysis identified two genes, PARL and BCL2L1, from blood tissues and three genes, ATG13, TOMM22, and SPATA33, from brain tissues as causal candidate genes associated with AD. The analysis pinpointed the possible epigenetic mechanisms, where specific methylation sites regulate the expression of these genes, potentially contributing to their association with AD. All findings were successfully replicated in UK Biobank cohorts.

**Conclusions:** The study emphasized the putatively causal relationships of mitophagy-related gene with AD. These underlying pathogenic mechanisms could pave the way for new approaches in early detection and therapeutic intervention for AD.

## Background

Alzheimer’s disease (AD), the leading cause of dementia in older adults worldwide, is a progressive, age-related neurodegenerative disorder characterized by memory impairment, cognitive decline, personality changes, and language disorders [1]. Its major pathological features include amyloid beta (Aβ) plaques, neurofibrillary tangles of hyperphosphorylated tau (p-tau), and neuroinflammation. Although the etiology of AD remains poorly understood, it is generally believed that the multifaceted interplay of genetic variation, environmental factors, and lifestyle underpins the pathogenesis of AD, especially in sporadic cases [2].

The brain is one of the most energy-demanding organs, and brain cells are particularly vulnerable to mitochondrial damage [3]. The homeostasis of both neurons and glial cells is heavily dependent on the normal function of mitophagy, the central component of mitochondrial quality control [4]. Mitophagy refers to the selective degradation of defective mitochondria via the autophagy pathway [5]. By removing dysfunctional mitochondria in neurons and supporting glial cells, mitophagy not only reduces cellular damage caused by reactive oxygen species (ROS) and mitochondrial DNA (mtDNA) leakage, but also enhances microglial phagocytosis of misfolded proteins like Aβ and tau in neurodegenerative diseases [6]. This process helps to attenuate neuroinflammation, primarily induced by the release of pro-inflammatory cytokines or ROS, and maintain cellular homeostasis in AD models, thereby preventing bioenergetic failure and cell death [7].

Mounting evidence implicates that mitophagy is significantly dysregulated in the brains of AD patients and AD models, which may be one of the underlying contributors to the manifestation and pathophysiology of the disease [8]. Altered expression levels of several mitophagy-related proteins, such as PINK1, p62, and LC3, have been observed in post-mortem human AD brains [9, 10].

Impaired mitophagy exacerbates oxidative damage and cellular energy depletion, leading to the aggregation of Aβ and p-tau proteins, which in turn disrupt the mitophagy mechanism itself [11]. PINK1/Parkin signaling, involving proteins such as PINK1, Parkin, and p62, is an important pathway regulating mitophagy [12]. PINK1-deficient mAPP mice exhibited earlier and increased Aβ accumulation, mitochondrial dysfunction, and cognitive impairment [13], while knocking out the p62 gene accelerated p-tau aggregation and neurodegeneration in PS19 mice [14]. Experimental evidence showed that upregulation of mitophagy-related proteins by genetic or pharmacological intervention could potentially increase mitophagy flux and ameliorate cognitive deficits in AD models [11, 13, 15].

Epigenetic modifications, particularly aberrant DNA methylation (DNAm) at CpG sites introduced by environmental factors, also play a vital role in AD development [16]. Both global and site-specific DNA hypomethylation have been found in post-mortem AD human brains and AD animal models [17, 18]. DNA methylation status in specific gene promoter or enhancer regions can affect Aβ deposition and neurofibrillary tangle formation by altering gene expression [19]. For example, hypomethylation in the promoter of BACE1 or PSEN1, both integral to the Aβ production, increases Aβ aggregates in AD mice by upregulating the expression of these genes [20, 21].

Multiple genome-wide association studies (GWAS) in AD patients have successfully identified several genetic susceptibility loci related to mitochondrial function, including APOE, TOMM40, and CLU, especially in late-onset AD cases [22–24]. Furthermore, methylome-wide association studies (MWAS) have also detected numerous AD-related DNA methylation marks, notably in genes such as APOE, MAPT, ANK1, and HOXA3 [25–27].

Despite these findings, which have expanded our knowledge of the genetic architecture and epigenetic mechanisms in AD, the associations identified from GWAS and MWAS may not be the direct genetic causes of the disease due to confounding effects from environmental and genetic factors [28, 29]. The specific role of mitophagy-related genes and their regulatory elements in AD remains elusive.

Summary data-based Mendelian randomization (SMR) is a method that combines GWAS data with molecular trait data, such as expression quantitative trait loci (eQTL) and DNA methylation QTL (mQTL), with the objective of investigating putative causal relationships between the molecular signatures and diseases [30]. Given that AD is a progressive neurodegenerative disorder affecting the central nervous system [31], analyzing the putative effects of genetic variants on brain-expressed genes may yield more meaningful insights than blood-based analyses. Accordingly, we employed a three-step SMR approach, integrating AD GWAS summary statistics with eQTL/mQTL data from blood and brain tissues separately, to investigate gene expressions and DNAm sites potentially causally linked to AD.

Furthermore, the replication analysis using data from UK Biobank datasets validated findings from the primary analysis to ensure the robustness and reliability of the discovery.

## Methods

### Study design

The study design is illustrated in Figure 1. Initially, a meta-analysis of transcriptomic data was conducted to recognize differentially expressed genes (DEGs) associated with mitophagy in AD patients. Subsequently, the three-step SMR approach integrated AD GWAS summary statistics with eQTL/mQTL data of DEGs to identify potential causal relationships between these genes and AD. The SMR tests were conducted using eQTL and mQTL data from blood tissues, separately. Genes and methylation sites identified as significant in the primary analysis were then validated using data from UK Biobank.

**Fig 1.**
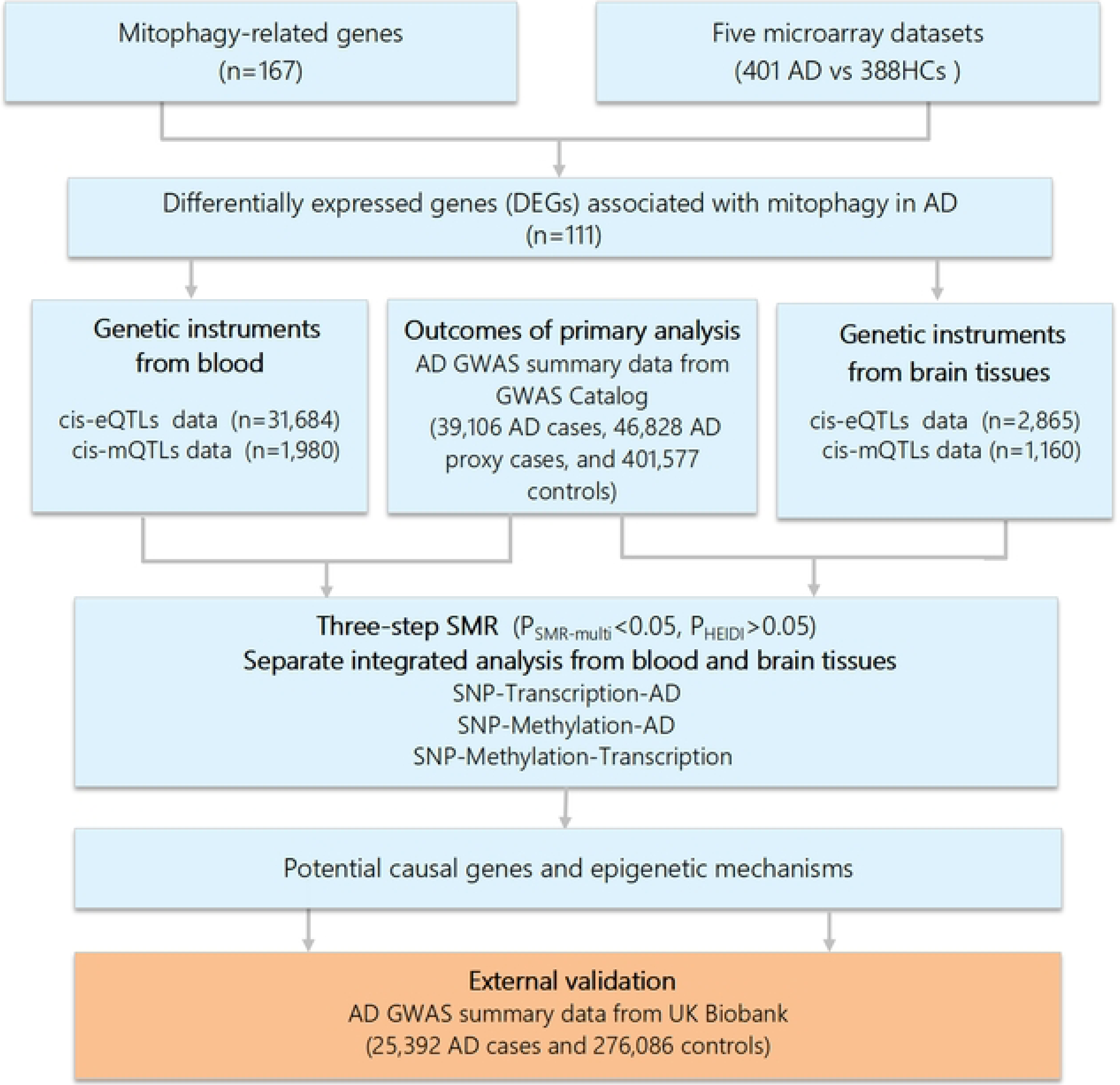
Flowchart of the study. AD, Alzheimer’s disease; cis-eQTLs, cis-expression quantitative trait loci; cis-mQTL, cis-methylation quantitative trait loci; GWAS, Genome-wide association studies; SMR, summary-based Mendelian randomization; SNP, single nucleotide polymorphisms.

### Data resources

Mitophagy-related genes were retrieved from the Gene Set Enrichment Analysis (GSEA), Kyoto Encyclopedia of Genes and Genomes (KEGG) and Gene Ontology Resource (GO) using the search terms “mitophagy”. Five microarray datasets for AD patients were gained from the Gene Expression Omnibus (GEO) database (http://www.ncbi.nlm.nih.gov/geo/) (S1 Table) [32–44].

In the primary analysis, GWAS summary statistics for AD from the GWAS Catalog were used, consisting of a total of 487,511 individuals, including 39,106 clinically diagnosed AD cases, 46,828 proxy cases, and 401,577 controls [45]. These data were obtained from a meta-analysis of AD GWAS datasets collected across 15 European countries, combining results from the European Alzheimer & Dementia Biobank (EADB) consortium and the UK Biobank. The data can be accessed publicly via the GWAS Catalog -GCST90027158 (https://www.ebi.ac.uk/gwas/studies/GCST90027158).

For external validation, we utilized GWAS summary data from the UK Biobank, encompassing a total of 301,478 individuals, with 25,392 AD cases and 276,086 controls [45]. The proxy-AD cases were determined based on questionnaire data in which individuals were asked if their parents had a diagnosis of dementia. All individuals analyzed in both the primary analysis and validation stages were of European ancestry.

Blood eQTL summary data of mitophagy-related genes were procured from eQTLGen, which contains statistics on blood gene expression from 31,684 individuals [46]. Blood mQTL data obtained through a meta-analysis implemented on two cohorts (n=1,980) [47]. Brain eQTL data were derived from BrainMeta v2 cis-eQTL summary data project (n= 2,865) [48], while the brain mQTL data were derived from the Brain-mMeta mQTL dataset (n = 1,160), generated through a meta-analysis that combined data from the studies by ROSMAP et al., Hannon et al., and Jaffe et al. [49]. This study focused on cis-eQTLs and cis-mQTLs, defined as single nucleotide polymorphisms (SNPs) located within a 1000 kb range upstream or downstream of the gene of interest.

For comprehensive details about the data used in the study, please refer to **Supporting information**.

### Statistical analysis

#### Identification of DEGs

DEGs related to mitophagy between AD patients and healthy controls (HCs) were identified using linear regression models for each dataset, with adjustments for age, sex, and other relevant covariates to account for potential confounding factors. A meta-analysis with a fixed-effects model was then carried out using the R package *metafor* to integrate the DEGs from individual datasets and derive the final set of DEGs [50].

#### SMR for detecting putative causal relationship

SMR tools were specifically designed to assess whether genetic variants influence complex traits through intermediate molecular traits, such as gene expression, DNA methylation, or protein abundance [30]. The 1000 Genomes European reference was used to calculate linkage disequilibrium [51], while the heterogeneity of dependent instruments (HEIDI) tests were employed for the assessment of heterogeneity [30].

The SMR analysis was conducted three times respectively using molecular QTL data from blood and brain tissues, with SNPs serving as genetic instrument variables in all three steps: (1) blood or brain eQTLs as exposures, with AD as the outcome; (2) blood or brain mQTLs as exposures, with AD as the outcome; (3) significant findings from step 1 as exposures, and findings from step 2 as outcomes. The significant causal associations were identified in accordance with the following criteria: (1) passing all three-step SMR with the threshold of *P_SMR-multi_* < 0.05; (2) demonstrating genome-wide significance, with *P* < 1×10⁻⁵ across all QTL datasets; and (3) displaying no significant heterogeneity (*P_HEIDI_* >0.05).

## Results

### DEGs associated with mitophagy

After removing duplicates, 167 unique mitophagy-related genes were collected from three databases (S2 Table). The recognition of DEGs was conducted on each of the five microarray datasets to contrast the expression levels of transcripts between AD patients (n=401) and HCs (n=388) (S1 Table). After the meta-analysis of results from these five datasets, 111 mitophagy-related genes were recognized as DEGs in AD (*P* < 0.05) (Fig 2 and S3 Table).

**Fig 2.**
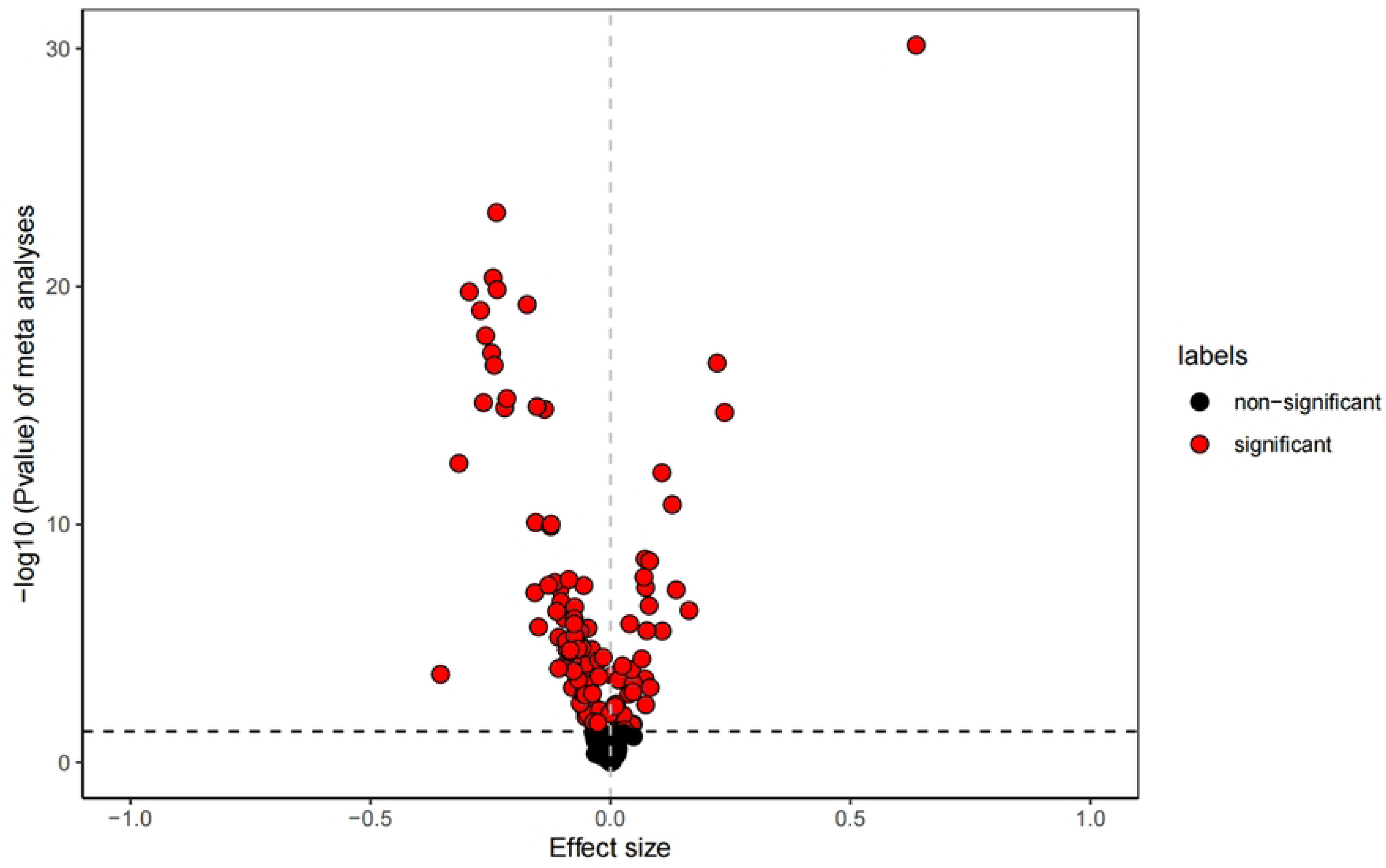
Volcano Plot of differentially expressed genes (DEGs) related to mitophagy in AD. A meta-analysis of five microarray datasets was conducted to compare gene expression between AD patients and healthy controls. In volcano plot, the x-axis represents meta-analysis effect sizes, while the y-axis shows the −log of meta *p* value, indicating statistical significance. Red dots highlight the 111 significant differentially expressed genes (DEGs), while black dots represent non-significant genes. The dashed line marks the significance threshold at p < 0.05.

### Integration of GWAS and mitophagy-related eQTL/mQTL data from blood and brain tissues

As outlined previously, we sought to identify candidate causal genes for AD and elucidate the epigenetic mechanisms involved. Our three-step SMR analysis combined AD GWAS summary statistics from the GWAS Catalog with blood or brain cis-eQTLs/cis-mQTLs data of 111 mitophagy-related DEGs.

Integrating blood eQTL data from the eQTLGen Consortium (n = 31,684) with AD GWAS summary data identified 5 mitophagy-related genes (*P_SMR-multi_* <0.05, *P_HEIDI_* >0.05) (S4 Table). Similarly, the integration of blood mQTL data (n = 1,980) with the same AD GWAS data identified 676 CpG sites (*P_SMR-multi_* <0.05, *P_HEIDI_* >0.05) (S5 Table). Further analysis combining the putative causal signals from the first two steps resulted in the identification of 6 CpG sites associated with the expression of two genes: PARL and BCL2L1 (*P_SMR-multi_* <0.05, *P_HEIDI_* >0.05) (S6 Table).

In a similar manner, leveraging eQTL (n = 2,865) and mQTL (n = 1,160) data from brain tissues along with the same AD GWAS summary statistics, we identified 9 mitophagy-related genes and 83 CpG sites (*P_SMR-multi_* <0.05, *P_HEIDI_* >0.05) (S7 and S8 Table). By combining findings from transcriptome and methylome data, the third analysis filtered out 7 CpG sites that putatively regulate the expression of three genes: ATG13, TOMM22, and SPATA33 (*P_SMR-multi_* <0.05, *P_HEIDI_*>0.05) (S9 Table).

The integration of multi-omics data from blood and brain tissues provides evidence supporting the causal relationships between mitophagy-related gene expression and AD pathogenesis, as shown in **Table 1**.

**Table 1.**
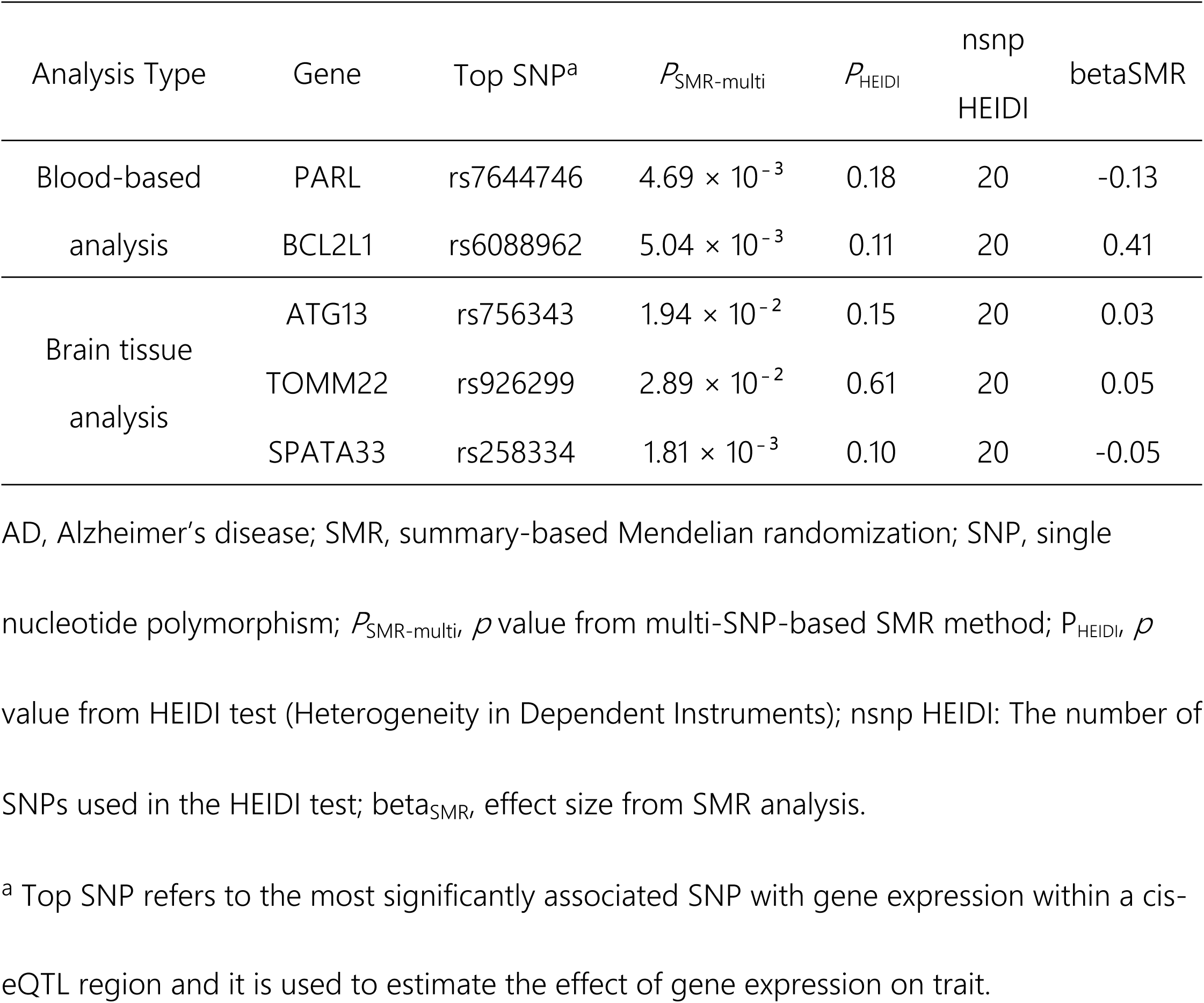
Associations of mitophagy-related gene expression with AD identified through three-step SMR analysis.

| Analysis Type | Gene | Top SNP<sup>a</sup> | $P_{SMR-multi}$ | $P_{HEIDI}$ | nsnp<br>HEIDI | betaSMR |
| --- | --- | --- | --- | --- | --- | --- |
| Blood-based<br>analysis | PARL | rs7644746 | $4.69 \times 10^{-3}$ | 0.18 | 20 | -0.13 |
| | BCL2L1 | rs6088962 | $5.04 \times 10^{-3}$ | 0.11 | 20 | 0.41 |
| Brain tissue<br>analysis | ATG13 | rs756343 | $1.94 \times 10^{-2}$ | 0.15 | 20 | 0.03 |
| | TOMM22 | rs926299 | $2.89 \times 10^{-2}$ | 0.61 | 20 | 0.05 |
| | SPATA33 | rs258334 | $1.81 \times 10^{-3}$ | 0.10 | 20 | -0.05 |

AD, Alzheimer's disease; SMR, summary-based Mendelian randomization; SNP, single nucleotide polymorphism; $P_{SMR-multi}$ , $p$ value from multi-SNP-based SMR method; $P_{HEIDI}$ , $p$ value from HEIDI test (Heterogeneity in Dependent Instruments); nsnp HEIDI: The number of SNPs used in the HEIDI test; beta<sub>SMR</sub>, effect size from SMR analysis.
<sup>a</sup> Top SNP refers to the most significantly associated SNP with gene expression within a cis-eQTL region and it is used to estimate the effect of gene expression on trait.

### Plausible epigenetic regulation of mitophagy-related genes by DNA methylation in AD

The three-step SMR analysis gave us an opportunity to speculate on the genetic regulation model of mitophagy-related genes in AD. One illustrative case is the PARL gene, which encodes a serine rhomboid protease located in the mitochondrial intermembrane space and is involved in regulating mitophagy by mediating the cleavage of PINK1 and PGAM5 [52, 53]. DNAm site cg22921096 in the 5’ untranslated region, 453 kbp downstream of PARL, was identified in the integration analysis of mQTL and AD GWAS summary statistics. The methylation level of this site exerted a positive effect on PARL expression (beta_SMR_ = 0.07) and a negative effect on AD (beta_SMR_ = -0.03). In addition, the PARL expression level was inversely related to the disease onset (beta_SMR_ = -0.13). Collectively, these results suggest that higher methylation level at this site upregulates the transcript level of PARL, potentially offering a protective effect against AD risk (Fig 3A).

**Fig 3.**
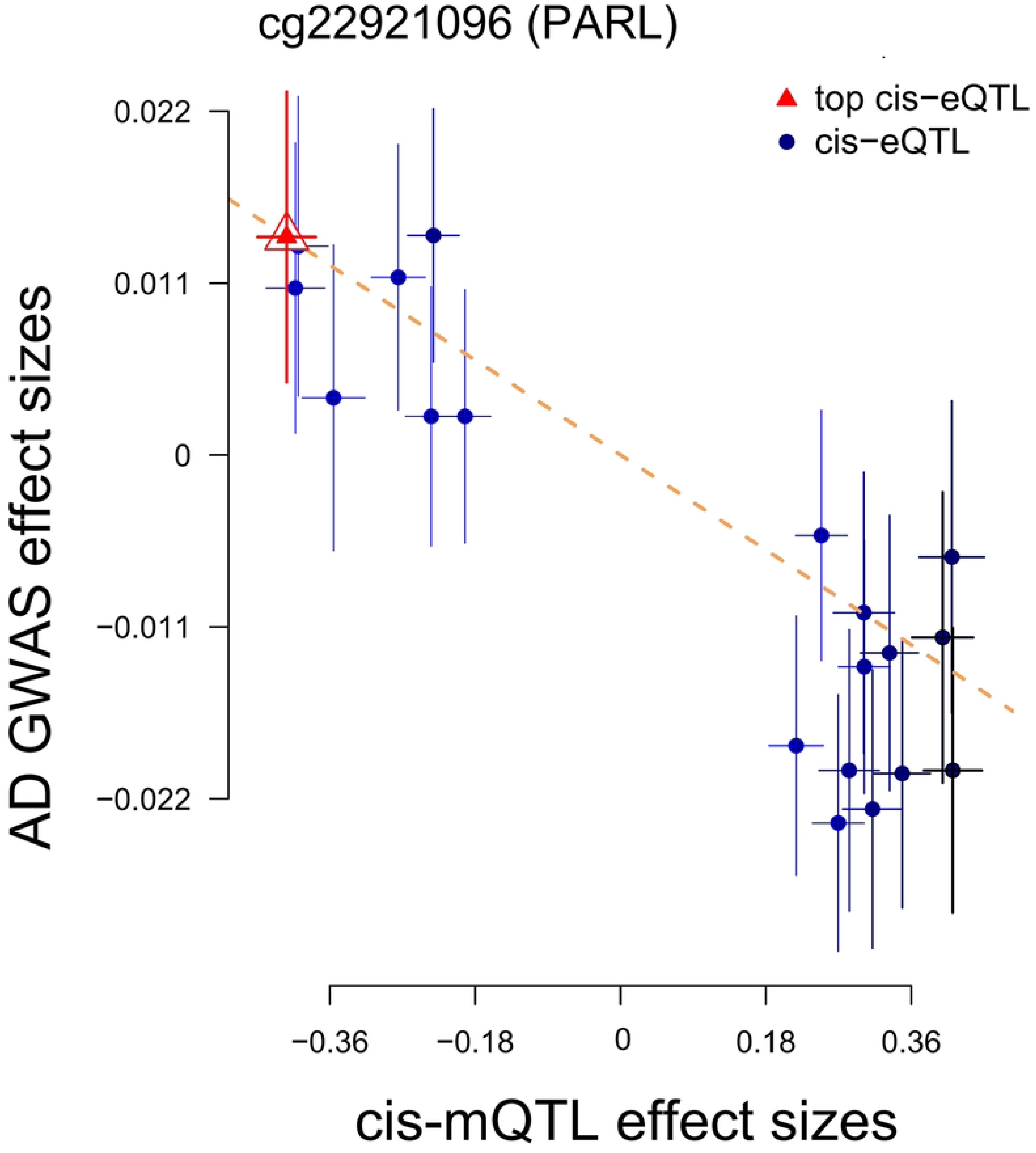
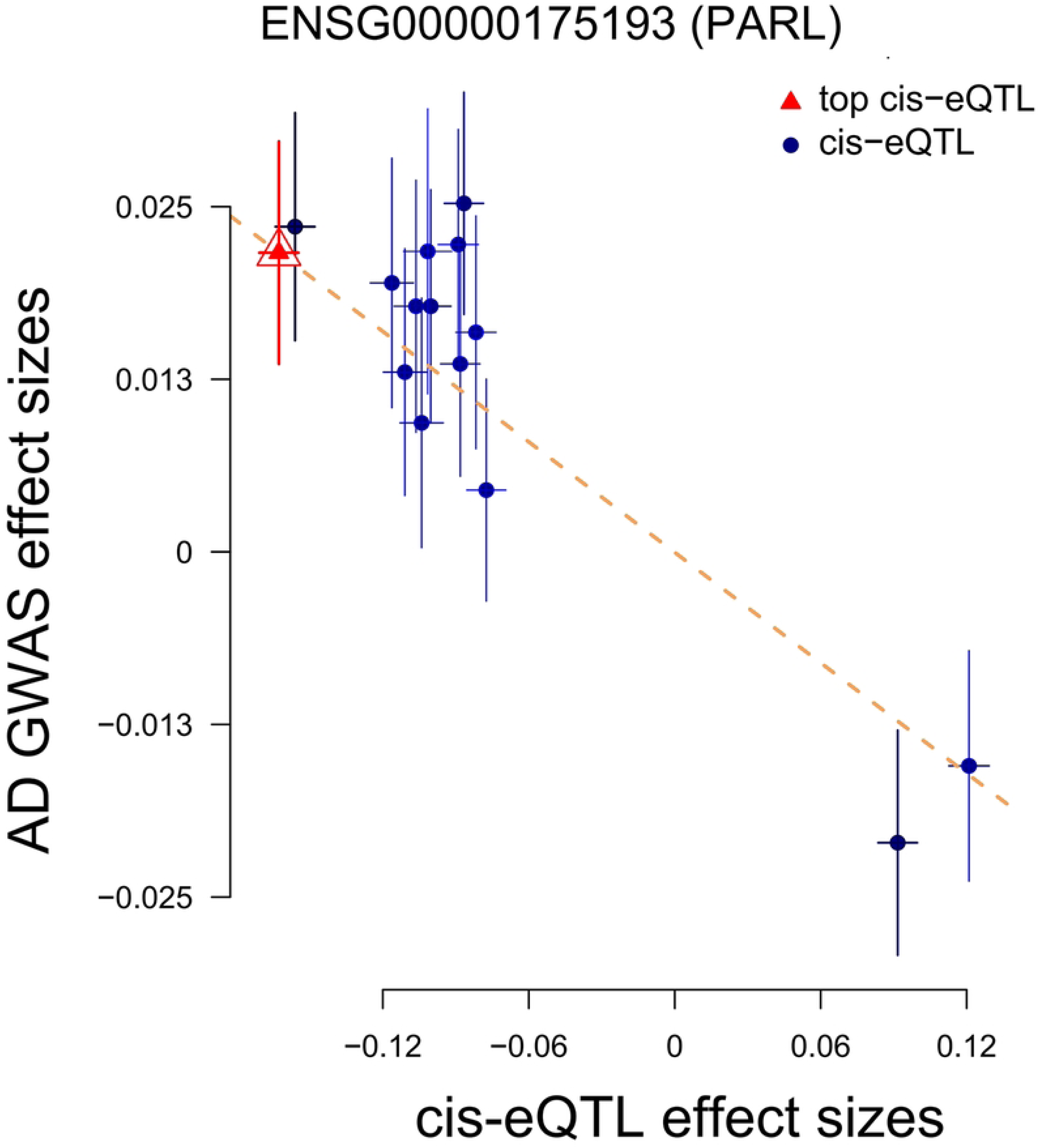
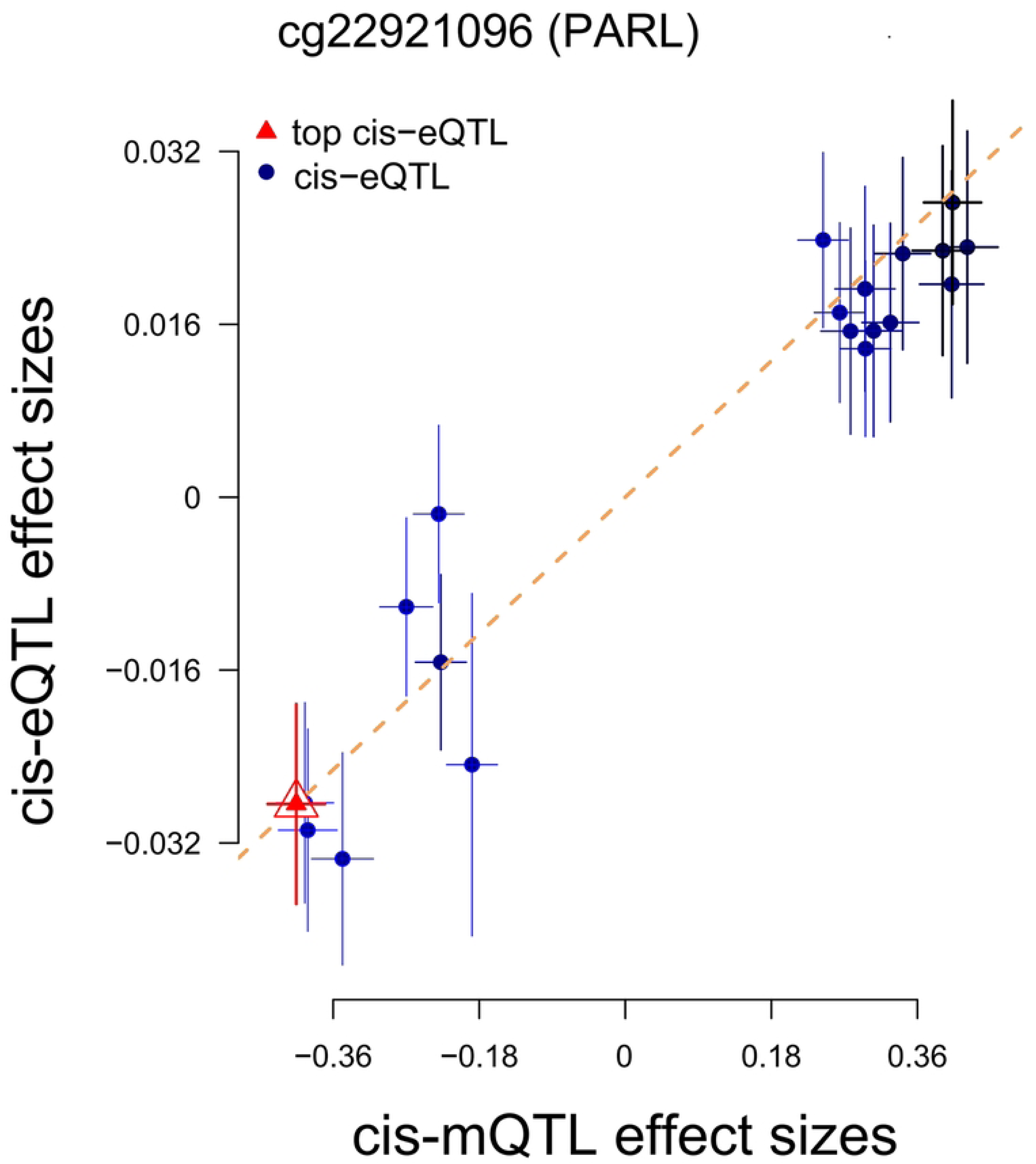
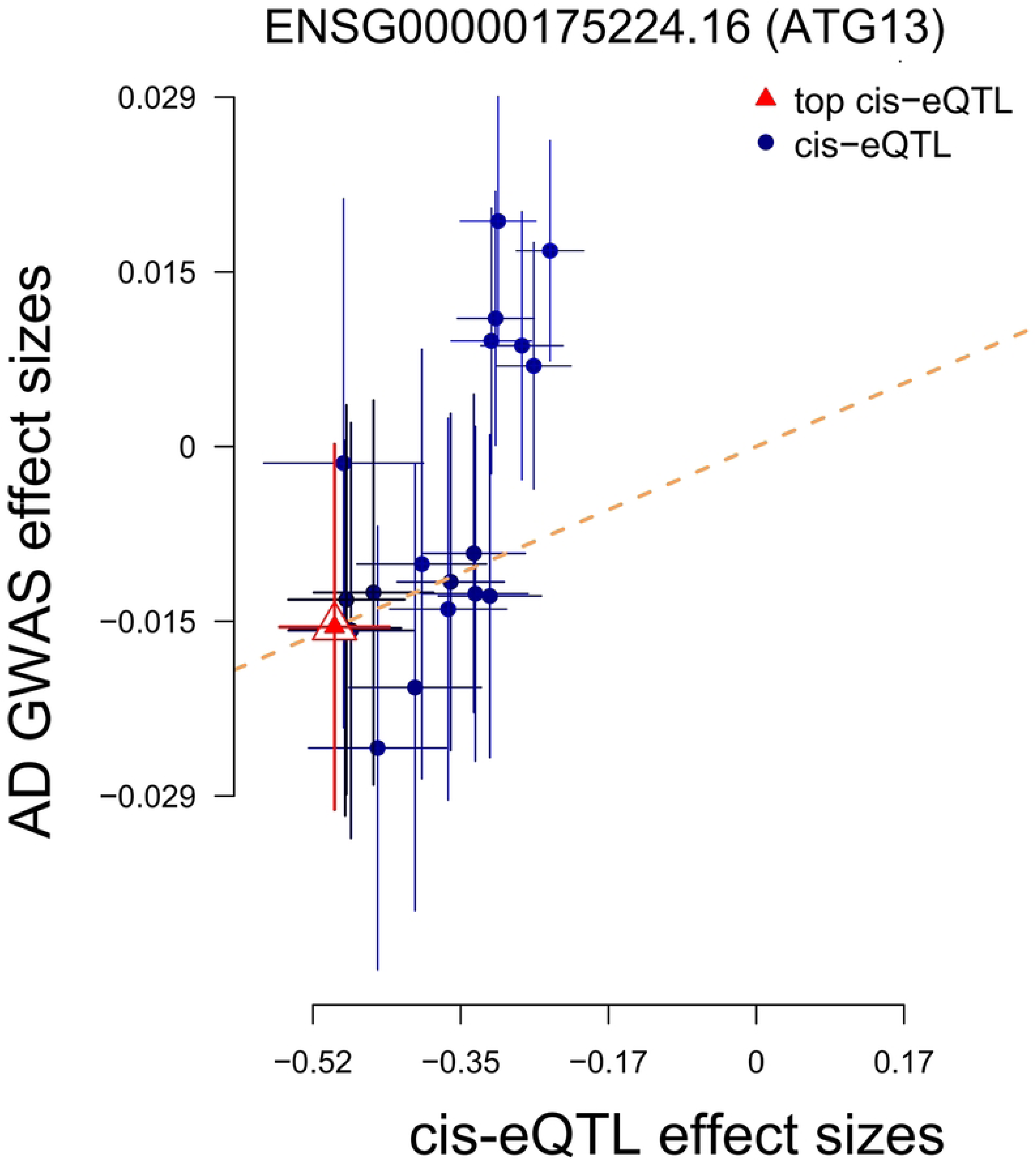
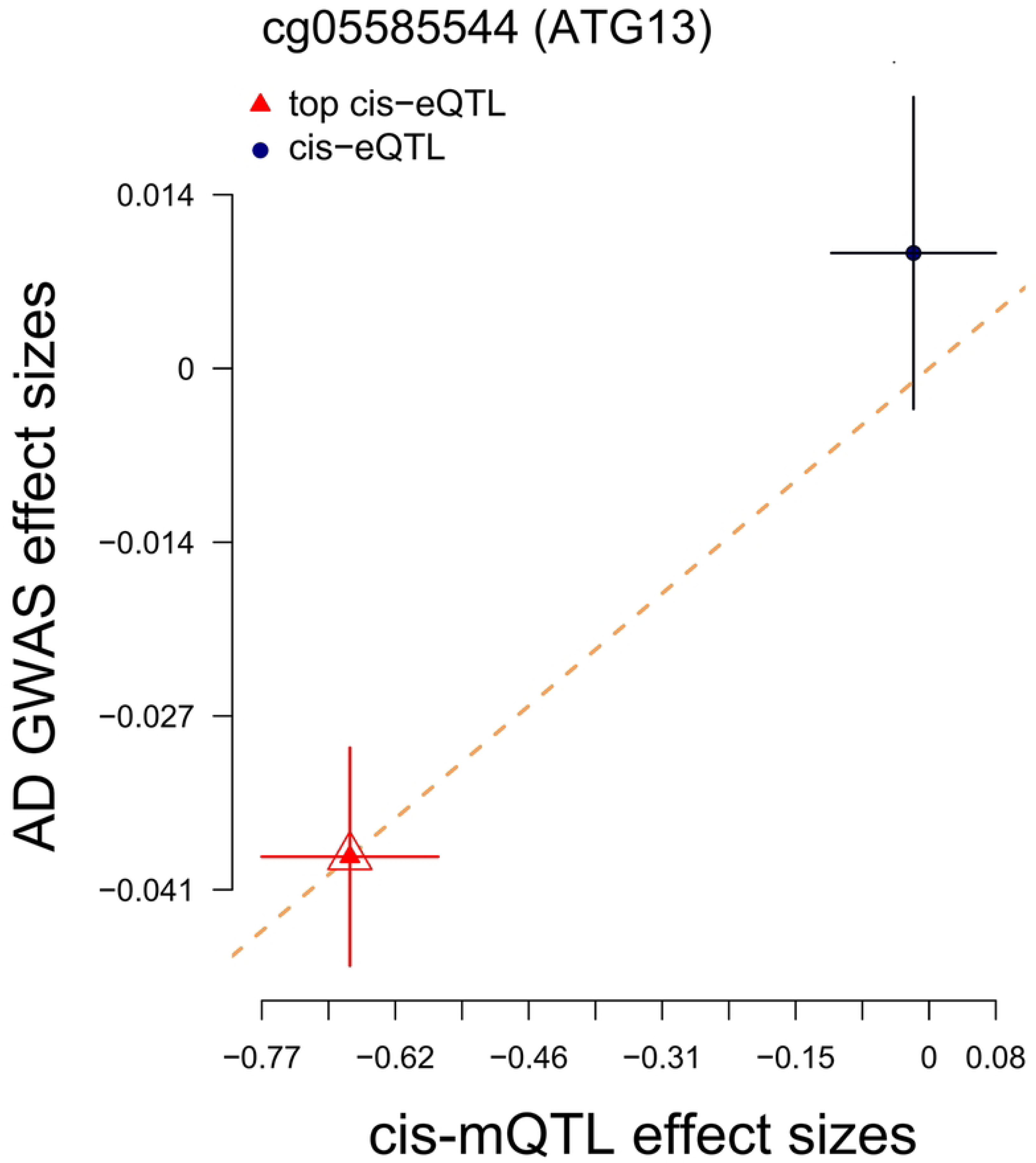
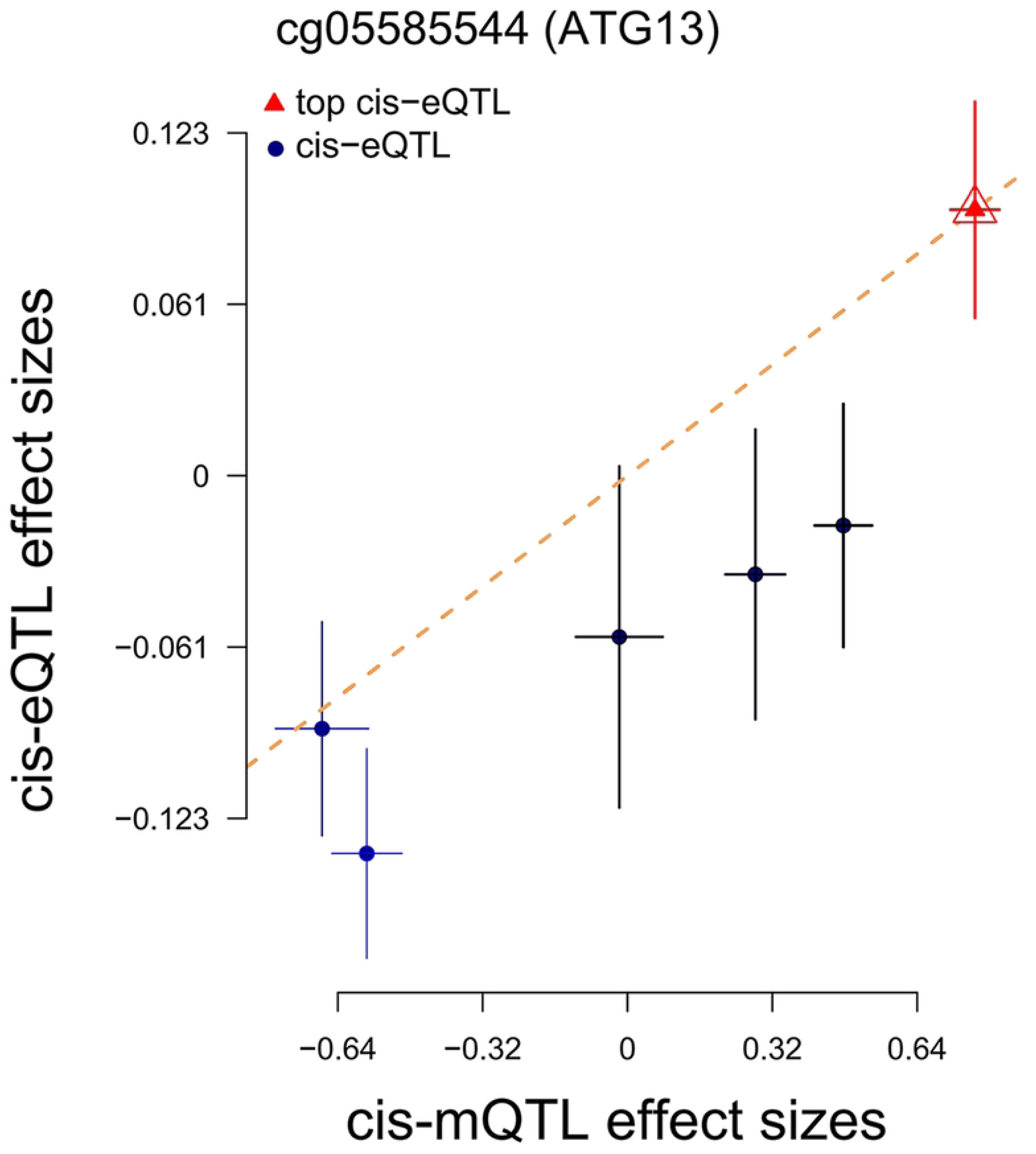
Three-step SMR analyses using molecular traits from blood and brain tissues. SMR tools were used to prioritize the mitophagy-related genes potentially causally associated with AD, along with relevant epigenetic mechanisms of genetic regulation (all *P_SMR-multi_* < 0.05, *P_HEIDI_* > 0.05). The panels, from left to right, depict the following analysis results: SMR between gene expression and AD GWAS on the left, SMR between gene methylation and AD GWAS in the middle, and SMR between gene methylation and gene expression. **Panels A** and **B** present the respective results for the genes PARL and ATG13. eQTL, expression quantitative trait loci; GWAS, Genome-wide association studies; mQTL, DNA methylation quantitative trait loci

ATG13, a key component of the ULK1 complex, is integral to the initiation of autophagy and mitophagy. Our analysis revealed a positive causal correlation between the ATG13 expression and DNAm site cg05585544, 957 kbp downstream of ATG13 (beta_SMR_ = 0.13). The elevated expression of ATG13 gene (beta_SMR_ = 0.03) and methylation levels of this CpG site (beta_SMR_ = 0.05) potentially increased the susceptibility to AD. Therefore, it is hypothesized that the genetic variation modulates the DNAm status at this CpG site, leading to upregulation of ATG13 expression, which could contribute to a heightened risk of AD (Fig 3B).

### External replication of mitophagy-related DEGs in AD GWAS summary statistics from UK Biobank

For external replication, we made use of AD GWAS data from the UK Biobank (n=361,141) and integrated them with eQTLs and mQTLs of previously detected genes to validate our primary findings. All tested genes and relevant DNAm sites showed significant associations with AD in the UK Biobank cohort, thereby reinforcing the validity of our primary research results (S10 Table).

## Discussion

This is the first study to identify mitophagy-related genes and their corresponding methylation sites potentially causally linked to AD. Dysregulation of mitophagy results in the buildup of defective mitochondria, which contribute to progression of AD pathologies such as Aβ aggregation, tau protein abnormalities, and neuroinflammation [5]. Targeting mitophagy has emerged as a promising therapeutic strategy, with animal model studies demonstrating its effectiveness in counteracting these pathological features and clinically improving cognitive deficits [54]. Since mitophagy-related DEGs in AD may either be a cause or an outcome of neurodegenerative processes [5, 55], it is essential to clarify the roles of these genes in AD development and their interactions with other biomolecules from a genetic perspective. By incorporating multi-omics data from different tissues through the SMR method, we identified a potentially causal relationship between five mitophagy-related genes and AD, and investigated their genetic regulatory mechanisms via epigenomic modifications. Among the genes filtered out by the three-step SMR, most of them have been characterized in AD field. For instance, PARL, a susceptibility locus for AD, regulates mitophagy through the PINK1-Parkin pathway [56]. A recent genome-wide survival study and gene analysis have reported that the downregulation of PARL expression correlated with worsening clinical symptoms of AD [57]. Lower PARL expression has been shown to increase p-tau levels in different AD models, while overexpression of PARL reduced this accumulation in basic experiment [57]. Our study further identified the CpG methylation in the 5’ untranslated region increased transcript level of PARL, potentially promoting its protective effect against AD (beta_SMR_ = -0.13).

Elevated levels of Bcl-xL have been observed in the brain tissues of AD patients [58]. The BCL2L1 gene encodes the Bcl-xL protein, an anti-apoptotic member of the Bcl-2 family, which modulates both apoptotic pathways and autophagy process, including mitophagy. Specifically, Bcl-xL inhibits mitophagy by binding to the BH3-like domain of Beclin 1, a key protein in the initiation and regulation of the process [59]. The inhibitory interaction between Beclin 1 and Bcl-xL could suppress activity of Beclin 1, thereby impairing mitophagy and exacerbating AD pathology [60].

ATG13, a vital component of the autophagy machinery, also plays an essential role in mitophagy processes [61]. At the onset of mitophagy, ATG13 forms the ULK complex with ULK1/2, ATG101 and FIP200, and the ULK complex initiates mitophagy by facilitating the formation and maturation of mitophagosomes [62], a structure that engulfs damaged mitochondria and delivers them to lysosomes for degradation.

Another significant finding from the three-step SMR analysis in brain tissue is the identification of TOMM22, a critical receptor within the translocase of the outer mitochondrial membrane (TOMM) complex, responsible for importing Parkin into mitochondria under normal conditions [63]. Previous studies have demonstrated that the destabilization of TOMM22 and TOMM40 acts as the switch to trigger the mitophagy [64]. Moreover, overexpression of TOMM22 partially inhibits Parkin-mediated mitochondrial clearance, suggesting that the proper regulation of TOMM22 is important for maintaining mitochondrial quality control [64]. Interestingly, SPATA33, identified as an autophagy mediator by recent experimental study, promotes the mitophagy in the male germline cells during spermatogenesis by interacting with the outer mitochondrial membrane protein VDAC2 and the autophagy-related protein ATG16L1 [65, 66]. The brain tissue analysis has suggested SPATA33 as a novel candidate gene potentially linked to AD development, although research in this area remains limited.

The strength of our investigation lies in the integration of multi-omics data from multiple tissues, providing a comprehensive analysis of the causal associations between mitophagy-related genes and AD. The three-step SMR with a more stringent statistical filtering criteria provided us stronger evidence to support our inference and enhanced our understanding of the genetic regulation mechanism involved in AD. Finally, all prioritized genes from the primary analysis were replicated in the other datasets, which further supported the reliability of the findings.

This study has some limitations that are worth acknowledging. First, eQTLs change in response to different cell states and environmental conditions [67]. Second, this study focused exclusively on cis-eQTLs and cis-mQTLs. Third, the participants included in these datasets were individuals of European ancestry, and the lack of ancestral diversity limits the applicability of our findings to more ethnically diverse populations.

## Conclusions

In conclusion, our study emphasized the potential causal links between mitophagy-related genes and AD. We identified biological mechanisms where DNA methylation modifications modulates the expression of specific mitophagy-related genes, thereby influencing the onset and progression of AD. These insights offer a deeper understanding of the pathogenesis of AD and hold significant implications for the development of novel therapeutic targets.

## Data Availability

All relevant data used in this study are publicly available. The GWAS summary statistics were obtained from the GWAS Catalog and the UK Biobank. The eQTL and mQTL summary data were from eQTLGen and yanglab. These datasets can be accessed at the following URLs: GWAS Catalog - GCST90027158: https://www.ebi.ac.uk/gwas/studies/GCST90027158. UK Biobank: https://broad-ukb-sumstats-us-east-1.s3.amazonaws.com/round2/additive-tsvs/AD.gwas.imputed_v3.both_sexes.tsv.bgz-OAD.gwas.imputed_v3.both_sexes.tsv.bgz, Blood eQTL summary data: https://www.eqtlgen.org/cis-eqtls.html, Blood and brain mQTL summary data, brain eQTL summary data: https://yanglab.westlake.edu.cn/software/smr/#DataResource. SMR tool? https://yanglab.westlake.edu.cn/software/smr/#Overview

https://www.eqtlgen.org/cis-eqtls.html

https://www.ukbiobank.ac.uk/

https://www.ebi.ac.uk/gwas/home

## Supporting information

**S1 Table. Characteristics of 5 microarray datasets.**

**S2 Table. 167 mitophagy-related genes list.**

**S3 Table. Meta-analysis of 111 differentially expressed genes related to mitophagy from five datasets (*p* <0.05).** Estimate, combined effect size estimate; SE, standard error of the effect size estimate; CI.lb, lower bound of the 95% confidence interval; CI.ub, upper bound of the 95% confidence interval.

**S4 Table. Summary-based Mendelian randomization (SMR) analysis from blood gene expression to AD (all *P_SMR-multi_* < 0.05, *P_HEIDI_* > 0.05).** ID, identifier; Chr, chromosome; bp, base pair position; SNP, single nucleotide polymorphisms; A1, effect (coded) allele; A2, other allele; Freq, frequency of effect allele; b, effect size; GWAS, Genome-Wide Association Studies; se, standard error; p, p-value; eQTL, expression quantitative trait locus; SMR, summary-based Mendelian randomization; SMR-multi, multi-SNP-based SMR; HEIDI, Heterogeneity in Dependent Instruments test; nsnp HEIDI, number of SNPs used in the HEIDI test.

**S5 Table. SMR analysis from blood DNA methylation to AD (all *P_SMR-multi_* < 0.05, *P_HEIDI_* > 0.05).**

SMR, summary-based Mendelian randomization.

**S6 Table. SMR analysis from blood DNA methylation to gene expression (all *P_SMR-multi_* < 0.05, *P_HEIDI_* > 0.05).** Expo, exposure; ID, identifier; Chr, chromosome; bp: base pair position; Outco; outcome; A1: effect allele; A2: other allele; Freq: frequency of the effect allele (A1); b: effect size; se: standard error; p, *p* value; SMR, summary-based Mendelian randomization; SMR-multi, multi-SNP-based SMR; HEIDI, Heterogeneity in Dependent Instruments test; nsnp HEIDI, number of SNPs used in the HEIDI test.

**S7 Table. SMR analysis from brain gene expression to AD (all *P_SMR-multi_* < 0.05, *P_HEIDI_* > 0.05).**

**S8 Table. SMR analysis from brain DNA methylation to AD (all *P_SMR-multi_* < 0.05, *P_HEIDI_* > 0.05).**

**S9 Table. SMR analysis from brain DNA methylation to gene expression (all *P_SMR-multi_* < 0.05, *P_HEIDI_* > 0.05).**

**S10 Table. External validation of the UK Biobank database**

## Acknowledgments

We thank all participants and investigators of the GWAS Catalog and UK Biobank.

## Author Contributions

Conceptualization: Weijian Zhang, Wenjia Chen, Hui Li.

Data curation: Weijian Zhang, Zhendong Guo.

Formal analysis: Weijian Zhang, Wenjia Chen.

Methodology: Weijian Zhang, Zhendong Guo.

Software: Weijian Zhang, Zhendong Guo.

Validation: Zhendong Guo.

Visualization: Weijian Zhang.

Writing – original draft: Weijian Zhang, Wenjia Chen, Zhendong Guo.

Writing – review & editing: Weijian Zhang.

Funding acquisition: Hui Li.

